# Effects of a 500-day LPG cookstove and fuel intervention delivered during gestation and infancy on height-for-age (HAZ) and stunting at ages 2, 3, 4 and 5 years: findings from a follow-up study of the multi-country HAPIN trial

**DOI:** 10.64898/2026.10.16.26365712

**Authors:** Md Abul Kalam, Sheela S. Sinharoy, Kyle Steenland, Lisa M. Thompson, Anaite Diaz-Artiga, John P. McCracken, Kalpana Balakrishnan, Vigneswari Aravindalochanan, Sarada Garg, Ghislaine Rosa, Florien Ndagijimana, Shirin Jabbarzadeh, Michael Johnson, Ajay Pillarisetti, Katie Kearns, Amy E. Lovvorn, Usha Ramakrishnan, Jennifer Peel, Thomas F Clasen, HAPIN Investigators

## Abstract

**Objective:** Reducing household air pollution exposure during gestation and infancy may have longer-term benefits for child linear growth. We examined whether a liquefied petroleum gas (LPG) cookstove and fuel intervention delivered during gestation and infancy could improve height-for-age Z scores (HAZ) and reduce the risk of stunting at ages 2–5 years in India, Rwanda, and Guatemala.

**Materials and method:** The Household Air Pollution Intervention Network (HAPIN) was a randomized controlled trial that enrolled ∼3200 pregnant women across Guatemala, India, Peru, and Rwanda. Women in the intervention group received an LPG cookstove and free fuel during pregnancy and until the infant reached 12 months of age. Following the HAPIN trial, we continued following children in Guatemala, India, and Rwanda (N=2169) and measured child height at ages 2, 3, 4, and 5 years (HAPIN 2.0). As prespecified in the study protocol, we used mixed-effects models, as well as linear and log-binomial regression, within the intention to treat (ITT) framework, to evaluate the intervention’s effect on HAZ and stunting. Subgroup analyses were performed to assess effect modification by child sex, maternal height, and intervention country.

**Results:** The intervention reduced gestational exposures to particulate matter (PM_2.5_: 80.7 vs 117.5 µg/m³), carbon monoxide (CO: 1.3 vs 2.0 ppm), and black carbon (BC: 8.6 vs 12.1 µg/m³) and infancy exposures (PM_2.5_: 36.9 vs 113.2 µg/m³; CO: 0.60 vs 2.2 ppm). Post-trial PM_2.5_ exposure were similar between arms: 99.9 vs 98.7 µg/m³. We found no statistically significant effect of the intervention on repeated measures of HAZ (β = -0.06, 95% CI: -0.16, 0.03 p=0.204), annual HAZ, stunting, nor severe stunting at any age during the follow-up. There was no statistically significant effect modification by subgroups.

**Conclusion:** Despite substantial reductions in air pollution exposures during the trial, the LPG intervention did not improve growth or reduce stunting risk from ages 2 through 5 years in this multi-country cohort. These findings, consistent with prior results of the lack of effects on growth through 12 months, provide additional evidence that a clean cooking intervention alone may not be sufficient to address childhood stunting in low-resource settings. Future exposure-response analyses will supplement these ITT analyses.

**Trial registration:** ClinicalTrials.gov NCT02944682

**Author summary:** *Why was this study done?:*

- Household air pollution (HAP) from cooking with biomass fuels has been associated with impaired fetal and child growth in low-and middle-income countries (LMICs).
- Earlier trials of clean cooking interventions delivered during pregnancy and infancy showed reductions in exposure to HAP but had no impact on child stunting or height-for-age z-scores at 1 year of age
- It remains unknown whether an intervention with a liquefied petroleum gas (LPG) cookstove and clean fuel that substantially reduces household air pollution exposure during pregnancy and infancy has longer-term effects on growth and stunting in later childhood, such as at ages 2 to 5 years.
- did the researchers do and find?
- We conducted a follow-up assessment of children who had been enrolled in the Household Air Pollution Intervention Network (HAPIN), a randomized controlled trial of an LPG cooking intervention, and compared height-for-age, stunting, and severe stunting between the intervention and control groups from ages 2 to 5 years in Guatemala, India, and Rwanda.
- We found no statistically significant effects of the intervention on repeated measures of HAZ or on annual HAZ, stunting, or severe stunting at any age, and no statistically significant effect modification by subgroups.

*What do these findings mean?:*

- Reducing household air pollution during pregnancy and infancy alone may not be sufficient to have effects on child linear growth through 5 years of age.
- The importance of the timing and duration of HAP exposure reductions for later child growth remains uncertain, and future exposure-response analyses could supplement these ITT findings.
- The main limitation of this study is loss to follow-up; children with repeated outcome measurements at all follow-up visits from 24 through 60 months represented 74% of the initially enrolled cohort.

## Introduction

Stunting is a form of linear growth faltering, defined as a height-for-age z-score (HAZ) more than two standard deviations below the World Health Organization (WHO) child growth standards reference median for age and sex (1). It is a major public health concern in low-and middle-income countries (LMICs) (1). Child stunting is associated with both immediate and long-term adverse health outcomes, including increased risk of morbidity, mortality, and impaired cognitive development (2). Stunting is also associated with adverse economic outcomes: a narrative review of randomized controlled trials and quasi-experimental and observational studies from 21 low-, middle-, and high-income countries linking child stunting and economic outcomes shows that childhood stunting is negatively associated with subsequent productivity, occupational status, and wages (3). Worldwide, 148.1 million children under 5 years of age (22.2 %) are stunted, with around 70% living in LMICs, and nearly two out of five children with stunting live in South Asia while another two out of five lived in sub-Saharan Africa (1).

An ample body of evidence suggests that stunting begins early in life, often in utero, and increases for at least the first 2 years after birth (4). However, longitudinal studies show that some children who were stunted at early ages (from birth to 2 years), showed improvements in later ages (4, 5). Conversely, later-onset stunting has been observed in various longitudinal studies, highlighting the evolving nature of our understanding of growth faltering (6). Common and known risk factors (such as household wealth, sanitation, hygiene, food insecurity, lack of dietary diversity, maternal height, infections) for child stunting in developing countries are rooted in broader socio-economic and environmental conditions (7).

Household air pollution (HAP), arising principally from the use of biomass fuels like wood, dung, charcoal, and agricultural crop waste as fuels for cooking, has been identified as a risk factor for adverse health outcomes including growth faltering and stunting (8). Prenatal exposure to air pollution can impair intrauterine growth and neurodevelopment through cellular-level mechanisms involving inflammation, oxidative stress, and epigenetic changes (9–12). Exposure to HAP during pregnancy is a cause of fetal growth restriction and prematurity, which in turn are associated with suboptimal child cognitive and motor development later in life (13–15). Although cross-sectional studies show a link between air pollution and childhood stunting (9, 16–21), only two studies have tested interventions experimentally: the Household Air Pollution Intervention Network (HAPIN) randomized controlled trial in Guatemala, India, Peru, and Rwanda and the Ghana Randomized Air Pollution and Health Study (GRAPHS). Both were conducted with one of the primary outcomes being stunting (22, 23). Despite reducing personal exposures to black carbon (BC), carbon monoxide (CO) and PM_2.5_ exposure, neither HAPIN nor GRAPHS interventions reduced the risk of low birth weight or stunting at age one year (22, 24–26).

To date, there have been no studies of the effect of clean cookstove interventions on child HAZ and stunting in childhood after 12 months of age. This continued interest is supported by substantial evidence highlighting the importance of these later years in subsequent child development (27, 28). Our aim was to determine whether the effects of an intervention that substantially reduced exposure to measured pollutants during gestation and the first year of life impacted longer-term growth. Here we report the effect of HAPIN intervention on HAZ at ages 2, 3, 4, and 5 in three countries, comparing children randomized to the control and intervention arms using intention-to-treat (ITT) analysis.

## Materials and methods

### Study design, randomization and participants

The study population was drawn from the HAPIN randomized control trial of an LPG cookstove and fuel. The study design has been described in detail elsewhere (29). The trial recruited pregnant women (N=3195) of >9 to <20 weeks gestation confirmed by ultrasound who were non-smokers and were using biomass as primary cooking fuel in Guatemala, India, Peru, and Rwanda. After enrollment and baseline measurements, participants were randomly assigned to either the intervention or control groups (assigned in 1:1 ratio), stratified by site, and followed for approximately 18 months until their child turned one year old. Households in the intervention group received a free LPG stove, unlimited LPG fuel, and behavioral reinforcements for the follow-up period, while control group households continued using primarily biomass fuel for cooking. One pregnant woman was enrolled from each participating household. Participants were randomized to the intervention or control arm using a block-randomization sequence generated centrally by the HAPIN Data Management Core, with block sizes selected at random. Allocation was concealed until assignment through the use of sealed envelopes. At each study site, participants chose from six sequentially numbered envelopes presented by field staff according to the trial allocation protocol. Participants and field staff were aware of treatment assignment because masking was not feasible for the cookstove intervention. Investigators remained blinded to the study data, however, and the primary analyses were completed with treatment assignment masked. Control participants received compensation specific to each country to balance the economic benefit received by households provided with the intervention.

Primary outcomes of the HAPIN RCT were birthweight, stunting and severe pneumonia in infants, and systolic blood pressure among older adult women living in the same home as the pregnant participant. Regular assessments of fetal or child exposure and growth were conducted throughout pregnancy (baseline, 24-28 weeks gestation, and 32-36 weeks gestation) and when the child reached 3, 6, and 12 months of age. Adherence to the HAPIN intervention was high, with over 96% of women in the intervention group reporting exclusive LPG use during pregnancy and with 67% completely abandoning traditional stoves for cooking and 31.4% completely removing them. Among intervention households retaining traditional stoves, median use was 0.0% over a 134-day follow-up (30).

Following the completion of the trial, the LPG stove and fuel intervention was discontinued. Most intervention households resumed using biomass for cooking; control households in India were offered the LPG cookstove and 18 months of clean fuel post-trial, Guatemala used coupons redeemable for items from a catalog; and Rwanda offered a choice among an LPG package, a solar home system, or cash compensation (31). Most of the participants chose alternatives forms of compensation for participating in the study, including cash or other goods (Williams, personal communication). Except control group in India, accordingly, most study participants would have been cooking on biomass fuels after the child born during the HAPIN trial was beyond 12 months of age. To assess the longer-term effects of the trial intervention, consenting participants from Guatemala, India, and Rwanda were enrolled in an extended follow-up study (HAPIN 2.0). Resources were unavailable to support follow-up activities in Peru. The primary aims of the HAPIN 2.0 study were to evaluate the effects of the original HAPIN intervention on child linear growth and neurodevelopment through 5 years of age. HAPIN 2.0 tracked 2260 children, out of 2318 live births in the participating 3 sites. In this analysis, we report the effect of the intervention on child HAZ, stunting, and severe stunting at ages 2 to 5 years among children who had at least one valid height measurements during 2, 3, 4 and 5 years of age visits.

### Outcome measurement

The primary outcome for this analysis is height-for-age z-scores (HAZ) measured longitudinally from ages 2 to 5 years. HAZ was calculated using the WHO Child Growth Standards, which define HAZ as the number of standard deviations a child’s height is from the median height of a healthy reference population of the same age and sex. We used WHO Anthro Stata installation package to calculate HAZ for the children ages 24-59 months and WHO AnthroPlus Stata macro package to calculate HAZ for children at 60 months, as recommended by WHO (32). Secondary outcomes include stunting and severe stunting from ages 2 to 5 years.

Following the WHO cut-off, stunting is defined as a HAZ more than two standard deviations below the WHO child growth standards reference median for age and sex and severe stunting is defined as a HAZ more than three standard deviations below the WHO child growth standards reference median for age and sex (33). Trained members of the HAPIN study team collected standing height annually at 24, 36, 48, and 60 months of age, using a SECA 217 stadiometer with a fixed vertical backboard and an adjustable head piece (SECA GmbH & Co. KG, Hamburg, Germany). Evaluators measured the height twice to the nearest millimeter. If the 2 height measurements were differed by more than 0.7 cm, then a third height measurement was taken. Before each round of measurement, the evaluators received training and standardization practices by an expert evaluators.

### Exposure measurement

Measurements and data collection of air pollutants have been described elsewhere (34). Briefly, 24-hour personal exposures to PM_2.5_ (particles ≤2.5 μm in aerodynamic diameter) were measured using the Enhanced Children’s MicroPEM (ECM) device (RTI International) and to CO using the Lascar EL-USB-CO300 monitor (Lascar Electronics). Black carbon levels were derived from PM_2.5_ filter samples at baseline and during pregnancy from the mothers, but were not quantified for the reconstructed infant exposures or during the post-trial phase. Women wore the monitoring devices in a vest or apron during data collection. Measurements were taken from pregnant women at three stages: before randomization (9–19 weeks), mid-pregnancy (24–28 weeks), and late pregnancy (32–36 weeks). Infants were monitored at 3, 6, and 12 months of age, with exposure assessed indirectly using microenvironmental reconstruction leveraging Bluetooth transmitters placed on children’s clothing and receivers and ECM monitors placed on the mom and in commonly frequented microenvironments (like the kitchen and sleeping area). Here, we report time-weighted (weighted by time before and after stove delivery) personal exposures to PM_2.5_, CO, and black carbon from baseline to late pregnancy (during gestation) in the intervention group, while for the control group we report the average exposures to these pollutants. For infancy, we report the average exposures of PM_2.5_ and CO for the infants, presented separately for the intervention and control groups, during the first year of life. During the post-trial period, children’s personal PM_2.5_ exposures were assessed at age 2, 3, 4, and 6 years using a combination of Ultrasonic Personal Aerosol Sampler (UPAS) and ECM devices across countries. Technical evaluations identified concerns regarding the reliability and validity of UPAS measurements, including invalid measurements in Rwanda and India due to failures in particle size selection and large particle deposition on filters. Therefore, we used PM_2.5_ measurements from the 3-year visit, the only follow-up visits with valid ECM data available across all study sites, to characterize post-trial exposure.

### Statistical analyses

The data presented in this manuscript were analyzed according to a pre-specified statistical analysis plan that was developed and approved before access to the data was obtained. All analyses followed the intention-to-treat (ITT) principle. Models were adjusted for randomization strata and baseline covariates that differed between the intervention and control groups, including maternal education, nulliparity, household food insecurity, and household ownership of a bank account.

The details of the statistical analysis plan are described in the Supplementary File (S-box 1). For the primary outcome -repeated measurements of height-for-age z-score (HAZ) from ages 2 to 5 years – we used mixed-effects models to estimate the intervention effect over time. The models included fixed and random effects to account for within-child correlation arising from repeated HAZ measurements and between-child variability. To estimate intervention effects on annual HAZ, we fitted separate linear regression models for HAZ measured at ages 2, 3, 4, and 5 years as our secondary outcomes. For annual stunting, we fitted age-specific log-binomial regression models to estimate risk ratios (RRs) comparing the intervention and control groups, with separate models fitted at ages 2, 3, 4, and 5 years. The same approach was used for annual severe stunting. When log-binomial models failed to converge, logistic regression models were used to estimate intervention effects.

To assess potential heterogeneity in intervention effects, we conducted interaction analyses using mixed-effects models. Effect modification was examined by country, maternal height [ a categorical variable based on WHO’s recommendation on short stature (35)], child sex, birthweight [a categorical variable based on WHO recommendation on low birthweight (36)] and time with the intervention [ <18 weeks vs ≥18 weeks]. Model fits were assessed using Akaike Information Criterion (AIC) to compare models, with lower values indicating better fit. All analyses were carried out in Stata SE software version 16.1 (StataCorp).

### Ethics review and trial registration statement

The study protocol was reviewed and approved by institutional review boards (IRBs) or Ethics Committees at Emory University (00089799), Johns Hopkins University (00007403), Sri Ramachandra Institute of Higher Education and Research (Deemed University) (IEC-NI/16/JUL/54/49; IEC-NI/21/OCT/80/120) and the Indian Council of Medical Research–Health Ministry Screening Committee (5/8/4-30/(Env)/Indo-US/2016-NCD-I; 2021-8184), Universidad del Valle de Guatemala (146-08-2016) and the Guatemalan Ministry of Health National Ethics Committee (11-2016), the London School of Hygiene and Tropical Medicine (11664; 22970) and the Rwandan National Ethics Committee (853/RNEC/2016; 317/2017; 357/2018; 194/2019; 929/2020; 64/2021; 45/2022; 47/2023; 17/2024; 666/2024). Written informed consent was obtained from participants prior to enrollment in the original trial, and additional written informed consent was obtained prior to participation in the follow-up study. The trial is registered with ClinicalTrials.gov (Identifier NCT02944682; URL: https://clinicaltrials.gov/study/NCT02944682). The trial protocol (including planned primary and secondary outcomes), informed consent form, and primary outcome statistical analysis plans can be accessed at https://clinicaltrials.gov/study/NCT02944682. De-identified data associated with these analyses will be available through the Emory University Dataverse repository, linked to the DOI of this publication (https://doi.org/10.15139/S3/4WGVHC, Dataverse Link: https://dataverse.unc.edu/dataset.xhtml?persistentId=doi:10.15139/S3/4WGVHC. Other materials can be accessed upon request.

## Results

### Participants

Between May 2018 and February 2020, a total of 2397 pregnant women in Guatemala (n=800), India (n=799), and Rwanda (n=798) were randomized to an intervention group (n=1194) or to the control group (n=1203) (Figure 1). Following randomization and before delivery, 43 women in the intervention and 36 women in the control group exited the study due to pregnancy loss, voluntary withdrawal, withdrawal by trial, or moving out from the trial area. Of the 2397 women randomized, 2318 (96.7%) resulted in live births, 1151 in the intervention group and 1167 in the control group. Before the children reached 12 months of age, 80 participants discontinued follow-up due to voluntary withdrawal, moving out of the trial area, or death (43= intervention and 37= control).

**Figure 1:**
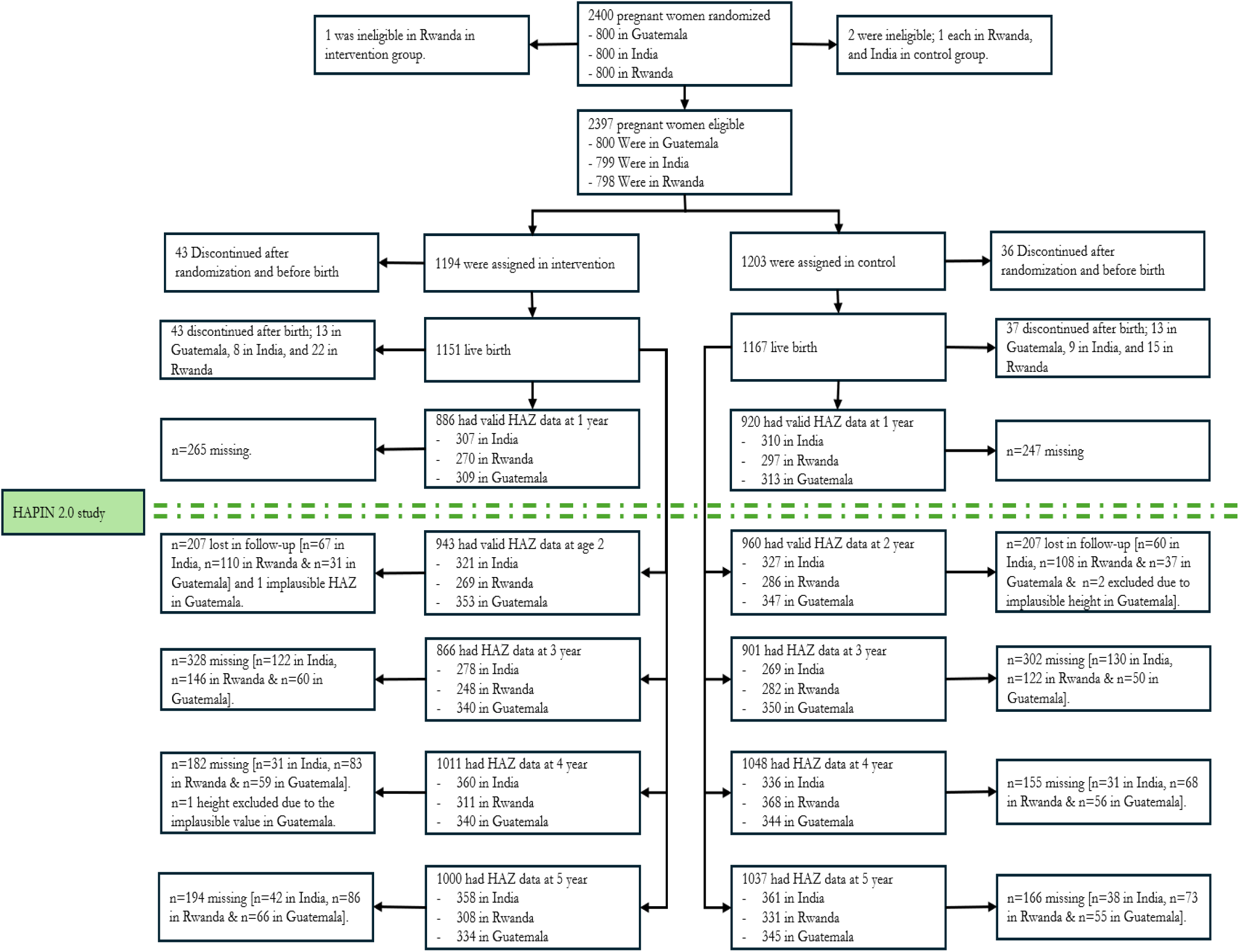
Participant Screening, Randomization, and Follow-up.

Valid height measurements were collected from 1903 at age 2 years (n=943 intervention and n=960 control), 1767 at age 3 years (n=866 intervention and n=901 control), 2059 at age 4 years (n=1011 intervention and n=1048 control) and 2037 at age 5 years (n=1000 in intervention and n=1037 control). This analysis is built on these 7766 height measurements. The number of children who completed all follow-up visits were 1610, with Guatemala contributing the largest sample while Rwanda the smallest (S-table 1). The number of children who participated in any follow-up visit were 2169, with India contributing the largest number of children (S-table 2). There were no substantive differences between intervention and control groups in the percentages of infants who were missing a height measurement (S-table 3).

**Table 1:** Maternal and household characteristics at baseline.

| Maternal characteristics at baseline | Intervention (n=1069) | Control (n=1100) | p-value* |
| --- | --- | --- | --- |
| Age (in years) | 25.2 ± 4.4 | 25.4 ± 4.5 | 0.509 |
| Average height (in centimeter) | 152.3 ± 6.6 | 152.8 ± 6.4 | 0.562 |
| Gestational age at baseline (in weeks) | 15.4 ± 3.0 | 15.2 ± 3.1 | 0.253 |
| Nulliparous - no | 638 (59.7%) | 708 (64.4%) | 0.025 |
| Dietary diversity | 3.0 ± 1.3 | 2.9 ± 1.3 | 0.398 |
| Education (in years) | 7.7 ± 3.9 | 7.3 ± 3.6 | 0.029 |
| Household characteristics at baseline |  |  |  |
| Bank account - no | 525 (49.1%) | 597 (54.3%) | 0.016 |
| Color television - no | 588 (55.0%) | 609 (55.4%) | 0.867 |
| Radio - no | 678 (63.4%) | 728 (66.2%) | 0.179 |
| Mobile phone - no | 161 (15.1%) | 178 (16.2%) | 0.472 |
| Household food insecurity score | 1.21 ± 2.0 | 1.50 ± 2.2 | 0.002 |
| Smoking in the household - no | 933 (87.4%) | 937 (85.3%) | 0.155 |
| Persons living in the household | 4.2 ± 2.1 | 4.2 ± 2.1 | 0.887 |
| For continuous variables, we report the mean, and standard deviation. For categorical variables, we report the count, percentage, and total number of observations. * p-values were generated through either t-tests or chi-square tests, depending on the type of variable. |  |  |  |

**Table 2:** Child characteristics at birth.

| Child characteristics at birth | Intervention (n=1069) | Control (n=1100) | p-value* |
| --- | --- | --- | --- |
| Sex - female | 512 (47.9%) | 528 (48.0%) | 0.961 |
| Preterm - no | 1011 (94.6%) | 1042 (94.7%) | 0.874 |
| Gestational age at birth (weeks) | 39.3 ± 1.6 | 39.3 ± 1.5 | 0.990 |
| Birth weight (kg) | 2825.1 ± 418.7 | 2824.5 ± 427.1 | 0.979 |
| For continuous variables, we report the mean, standard deviation, and total number of observations. For categorical variables, we report the count, percentage, and total number of observations. * p-values were generated through either t-tests or chi-square tests, depending on the type of variable. |  |  |  |

**Table 3:** Personal Exposures to Fine Particulate Matter, Black Carbon, and Carbon Monoxide.

| Pollutants<sup>‡</sup> | Intervention [mean, SD, (n)<sup>§</sup>] | Control [mean, SD, (n)<sup>§</sup>] |
| --- | --- | --- |
| **Average maternal exposures during gestation\*** |  |  |
| Fine particulate matter (µg/m<sup>3</sup>) | 80.7 ± 71.2 (992) | 117.5 ± 89.0 (1188) |
| Black carbon (µg/m<sup>3</sup>) | 8.6 ± 6.6 (886) | 12.1 ± 7.2 (1174) |
| Carbon monoxide (ppm) | 1.3 ± 6.6 (1049) | 2.0 ± 2.6 (1198) |
| **Average infancy (from birth to 12 months of age)# exposures** |  |  |
| Fine particulate matter (µg/m<sup>3</sup>)~ | 36.9 ± 32.9 (1098) | 113.2 ± 117.1 (1126) |
| Carbon monoxide (ppm)~ | 0.60 ± 1.1 (1112) | 2.2 ± 3.8 (1129) |

| Post trial exposure |  |  |
| --- | --- | --- |
| Fine particulate matter ($\mu\text{g}/\text{m}^3$) at age 3 | $99.9 \pm 140.7$ (510) | $98.7 \pm 108.1$ (533) |
| *These exposures were measured at Baseline, and two gestational visits and we report the time-weighted value here, with baseline weighted by time before stove delivery, the average of last two visits weighted by time after delivery. #These exposures were measured at age 3, 6 and 12 months of age by an indirect method (i.e., infants wore a beacon emitter, and we report the average exposures of these three months. ~ These exposures had missing measurements for infants at different visits; therefore, in the case of missing in infants exposures we used maternal exposures as proxies, as mother–child exposures to these pollutants are highly correlated ($r \approx 0.90$). § For all pollutants, we report mean, plus–minus values are means ±SD. ‡Personal exposures to fine particulate matter, black carbon, and carbon monoxide were measured over a period of 24 hours at baseline and after randomization. The abbreviation ppm denotes parts per million |  |  |

### Baseline characteristics

Table 1 describes the pregnant participants and household characteristics at baseline for our analytical sample (the children who participated in the follow-up; n=2169) where it shows that these characteristics were largely comparable between intervention and control groups. The mean maternal age was similar (25.2 vs. 25.4 years), as were the mean maternal height (152.3 vs. 152.8 cm), mean dietary diversity score (3.0 vs. 2.9), and mean gestational age at enrollment (15.4 vs. 15.2 weeks). Differences between arms were found for nulliparity (with 59.7% and 64.4% in respectively in intervention and control) and years of education (7.7 vs 7.3 years). Differences in household characteristics were modest but notable. Control households were more likely to lack a bank account (55.0% vs. 49.5%) and had a slightly higher household food insecurity score (1.51 vs. 1.3). Ownership of color televisions, radios, mobile phone, the number of people living in the household, and the absence of smoking in the household were similar between groups.

### Child characteristics

Infant characteristics at birth were very similar between the intervention and control groups (Table 2; the characteristics of missing children are given in S-table 4). The proportion of female infants was nearly identical (47.4% vs. 47.8%), indicating balanced sex distribution across groups. Preterm birth rates were also comparable, with the majority of births occurring at term in both groups (94.0% vs. 94.2%). Mean gestational age at birth did not differ between groups (39.3 weeks). Birth weight was likewise consistent (2823.9 vs 2823.2g). Overall, infant outcomes at birth showed no meaningful differences between groups. Dietary diversity scores were also similar between intervention and control groups at all ages, with a slight increase over time in both groups and no consistent intervention-related difference (S-table 5).

**Table 4:**
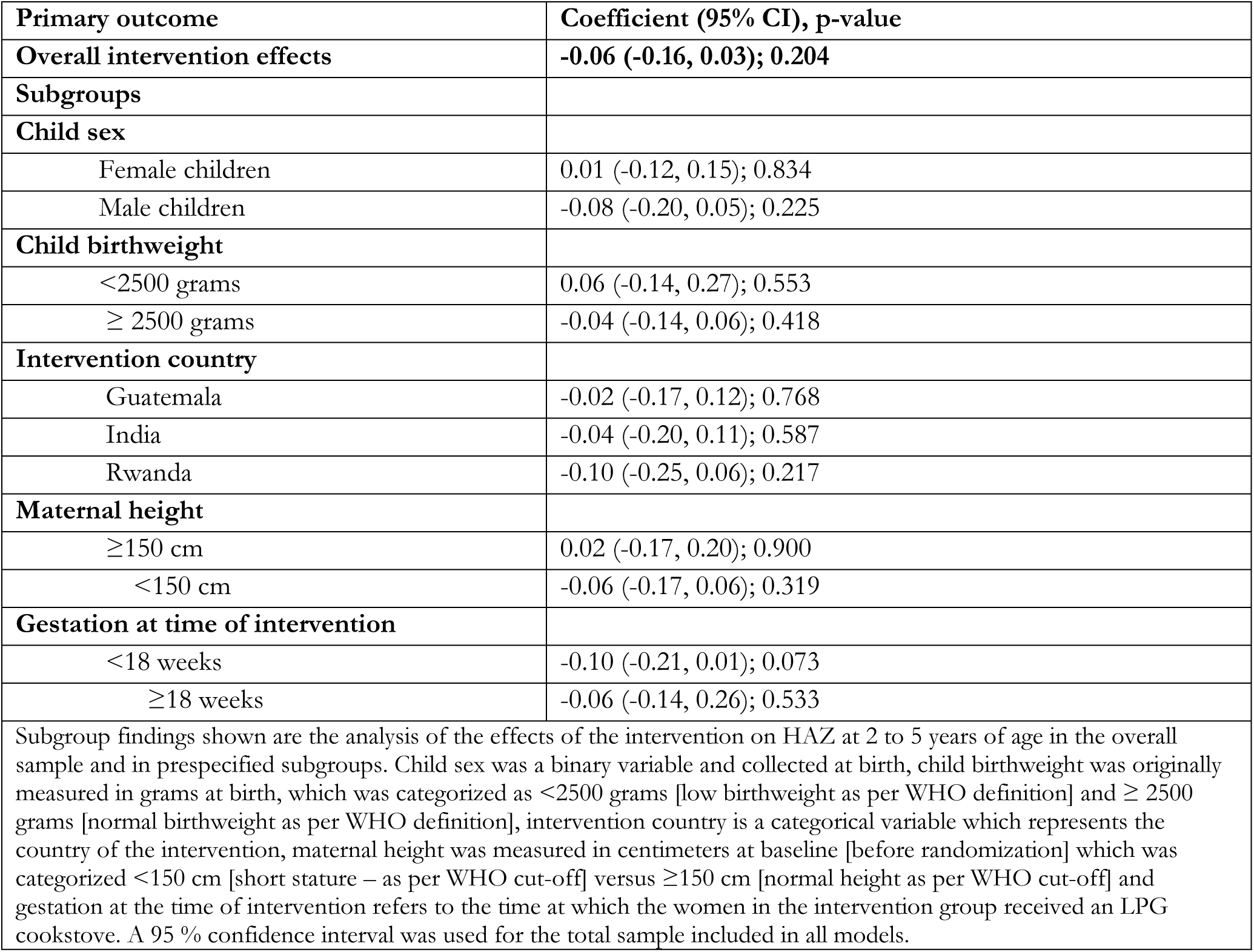
Findings from mixed-effect model on the effect of the intervention on HAZ.

| Primary outcome | Coefficient (95% CI), p-value |
| --- | --- |
| **Overall intervention effects** | **-0.06 (-0.16, 0.03); 0.204** |
| **Subgroups** |  |
| **Child sex** |  |
| Female children | 0.01 (-0.12, 0.15); 0.834 |
| Male children | -0.08 (-0.20, 0.05); 0.225 |
| **Child birthweight** |  |
| <2500 grams | 0.06 (-0.14, 0.27); 0.553 |
| ≥ 2500 grams | -0.04 (-0.14, 0.06); 0.418 |
| **Intervention country** |  |
| Guatemala | -0.02 (-0.17, 0.12); 0.768 |
| India | -0.04 (-0.20, 0.11); 0.587 |
| Rwanda | -0.10 (-0.25, 0.06); 0.217 |
| **Maternal height** |  |
| ≥150 cm | 0.02 (-0.17, 0.20); 0.900 |
| <150 cm | -0.06 (-0.17, 0.06); 0.319 |
| **Gestation at time of intervention** |  |
| <18 weeks | -0.10 (-0.21, 0.01); 0.073 |
| ≥18 weeks | -0.06 (-0.14, 0.26); 0.533 |
| Subgroup findings shown are the analysis of the effects of the intervention on HAZ at 2 to 5 years of age in the overall sample and in prespecified subgroups. Child sex was a binary variable and collected at birth, child birthweight was originally measured in grams at birth, which was categorized as <2500 grams [low birthweight as per WHO definition] and ≥ 2500 grams [normal birthweight as per WHO definition], intervention country is a categorical variable which represents the country of the intervention, maternal height was measured in centimeters at baseline [before randomization] which was categorized <150 cm [short stature – as per WHO cut-off] versus ≥150 cm [normal height as per WHO cut-off] and gestation at the time of intervention refers to the time at which the women in the intervention group received an LPG cookstove. A 95 % confidence interval was used for the total sample included in all models. |  |

**Table 5:** Effect of the intervention on stunting, severe stunting and HAZ at ages 2, 3, 4 and 5.

| Outcomes | Intervention | Control | Effect of the intervention |
| --- | --- | --- | --- |
| **At 2 years** |  |  |  |
| Height for age Z score* | -1.79 ± 1.12 (-5.95 - 1.69) | -1.77 ± 1.12 (-5.97 - 1.58) | -0.05 (-0.14, 0.05); 0.354 |
| Stunting# | 402 (42.6%) | 410 (42.7%) | 1.01 (0.92, 1.11); 0.829 |
| Severe stunting# | 133 (14.1%) | 122 (12.7%) | 1.09 (0.86, 1.38); 0.468 |
| **At 3 years** |  |  |  |
| Height for age Z score* | -1.80 ± 0.97 (-4.69 - 1.09) | -1.78 ± 0.99 (-5.96 - 1.28) | -0.06 (-0.15, 0.03); 0.210 |
| Stunting# | 359 (41.5%) | 363 (40.3%) | 1.09 (0.97, 1.23); 0.136 |
| Severe stunting# | 92 (10.6%) | 95 (10.5%) | 1.13 (0.84, 1.52); 0.422 |
| **At 4 years** |  |  |  |
| Height for age Z score* | -1.68 ± 0.98 (-4.55 - 1.2) | -1.66 ± 0.99 (-5.21 - 1.88) | -0.06 (-0.15, 0.02); 0.139 |
| Stunting# | 385 (38.1%) | 380 (36.3%) | 1.07 (0.95, 1.19); 0.279 |
| Severe stunting# | 81 (8.0%) | 87 (8.3%) | 1.02 (0.76, 1.39); 0.869 |
| **At 5 years** |  |  |  |
| Height for age Z score* | -1.48 ± 0.94 (-4.14 - 1.56) | -1.48 ± 0.95 (-5.00 - 2.02) | -0.02 (-0.11, 0.06); 0.584 |
| Stunting# | 290 (29.0%) | 302 (29.1%) | 1.02 (0.88, 1.17); 0.802 |
| Severe stunting# | 44 (4.4%) | 47 (4.5%) | 0.95 (0.62, 1.45); 0.808 |
| *Refers to the coefficient for the regression estimates for height for age z score outcome at age 2, 3, 4 and 5. # Refers to the risk ratio for the stunting and severe stunting outcomes. For continuous outcome (HAZ), we report mean, standard deviation and range and for binary outcomes (stunting and severe stunting), we report count, and percentage. We used linear regression, and log-linear regression models to estimate the effects of the intervention on annual HAZ, stunting and severe stunting respectively. A 95 % confidence interval was used for the total sample included in all models. |  |  |  |

### Exposure to air pollutants

During gestation, mothers in the intervention group experienced lower exposures to household air pollutants compared with those in the control group (Table 3). The average time-weighted exposure to fine particulate matter (PM_2.5_) was 80.7 μg/m³ among intervention participants, compared with 117.5 μg/m³ in controls. Black carbon exposure was similarly reduced (8.6 μg/m³ versus 12.1 μg/m³), as was carbon monoxide (1.3 ppm versus 2.0 ppm). These reductions persisted during infancy: infants in the intervention group had markedly lower exposures from birth through 12 months, with average PM_2.5_ levels of 36.9 μg/m³ compared with 113.2 μg/m³ in controls and carbon monoxide levels of 0.60 ppm versus 2.2 ppm. Overall, the intervention successfully reduced both maternal and infant exposure to measured pollutants during pregnancy and infancy. At age 3 years, mean post-trial PM_2.5_ exposure was similar between the intervention (99.9 ± 140.7 μg/m³; n=510) and control groups (98.7 ± 108.1 μg/m³; n=533).

### Effect of the intervention on primary and secondary outcomes

Figure 2 shows the mean height-for-age Z-score (HAZ) from age 2 to 5 years by trial arm, with 95% confidence intervals. Both the intervention and control groups show improvements in mean HAZ over time, with parallel upward trends. While the intervention group starts slightly lower, the patterns remain similar and overlapping, suggesting no substantial difference in mean HAZ between the groups across ages.

**Figure 2:**
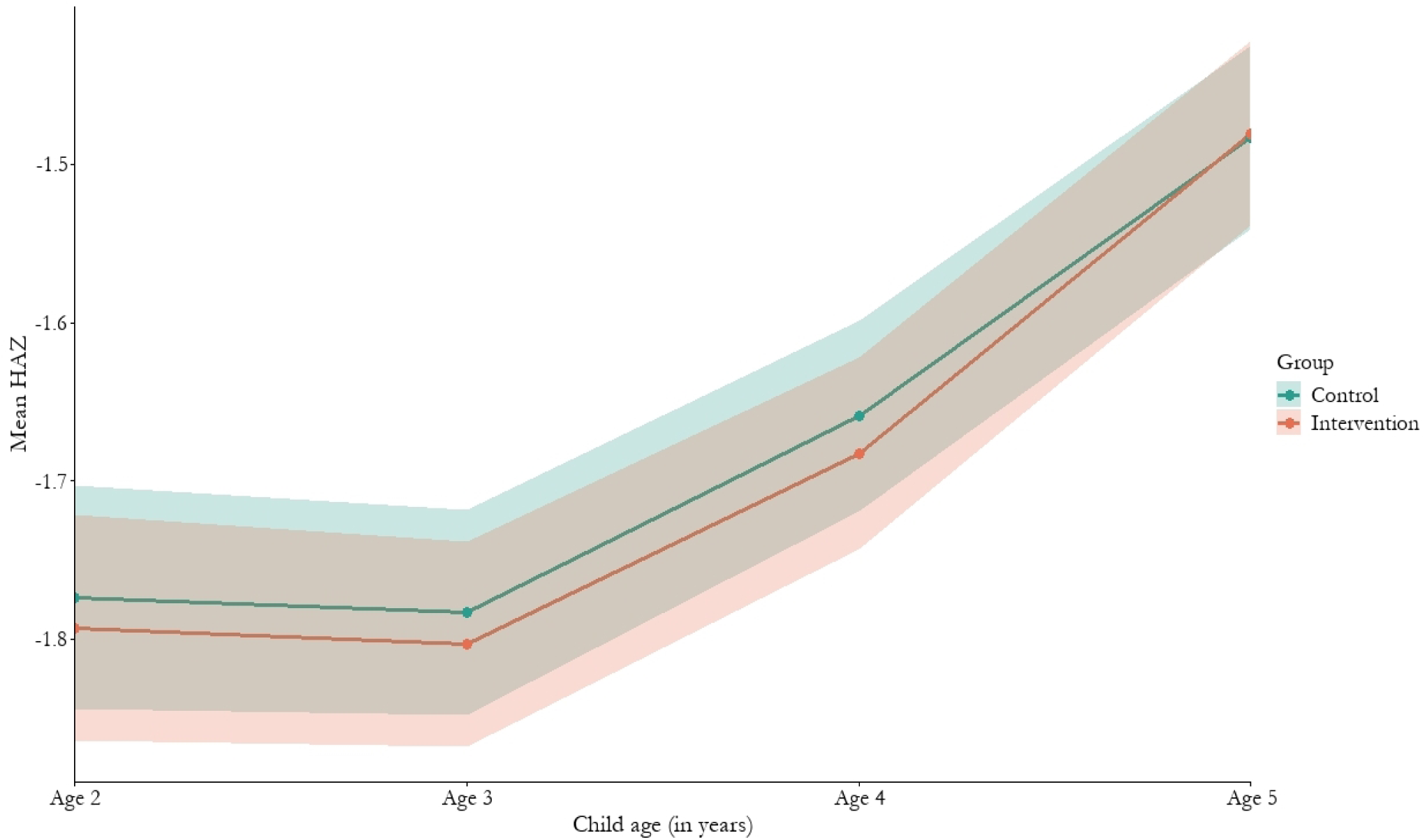
Mean height-for-age Z-score from age 2 to 5 years by trial arm, with 95% confidence intervals.

### Intervention effects on height-for-age z-scores (HAZ) repeated measures from ages 2 to 5 years

In the mixed-effect model for repeated measurements, height-for-age Z scores at 2, 3, 4, and 5 years of age were used (n = 7766 HAZ measurements). The mixed-effects model, shown , revealed no statistically significant difference in mean HAZ between the intervention and control groups from age 2 to age 5 years (*α*_1_= -0.06, 95% CI: -0.16, 0.03 p=0.204). There were also no differences in time trends between groups (Table 4). The main full mixed-effects model is described in supplementary file (S-table 6). Tests for interactions showed no statistically significant effect modification by subgroup, indicating that the intervention effect did not differ meaningfully across subgroups. These findings suggest that the intervention did not have an effect on HAZ.

### Intervention effects on annual HAZ, stunting, severe stunting, and HAZ

Table 5 outlines the annual results of intervention effects on HAZ, stunting, and severe stunting. Analyses from annual HAZ models showed that mean HAZ scores increased over time. In the intervention group, the mean HAZ scores were -1.79 at age 2, -1.80 at age 3, -1.68 at age 4, and -1.48 at age 5. Similarly, in the control group, the mean HAZ scores were -1.77 at age 2, -1.78 at age 3, -1.66 at age 4, and -1.48 at age 5. The annual linear regression models did not show any significant effect of the intervention on mean HAZ at any of the annual time points compared with the control group.

Analyses from annual stunting models showed similar null results (Table 5). For children born to women in the intervention group, the prevalence of stunting was 42.6% at age 2, 41.5% at age 3, 38.1% at age 4, and 29% at age 5. Similarly, in the control group, the prevalence of stunting was 42.7% at age 2, 40.3% at age 3, 36.3% at age 4, and 29.1% at age 5. The ITT analyses showed that the intervention did not reduce the risk of stunting at any of the annual time points compared with the control group. Analyses from annual severe stunting models were similarly null (Table 5). In the intervention group, the prevalence of severe stunting was 14.1% at age 2, 10.6% at age 3, 8.1% at age 4, and 4.4% at age 5. Similarly, in the control group, the prevalence of severe stunting was 12.7% at age 2, 10.5% at age 3, 6.3% at age 4, and 4.1% at age 5. In the subgroup analyses of annual HAZ (Figure 3.), no subgroup appeared to benefit from the intervention more than others at any time point in HAZ models (detail results are provided in the supplementary file (S-table 7).

**Figure 3:**
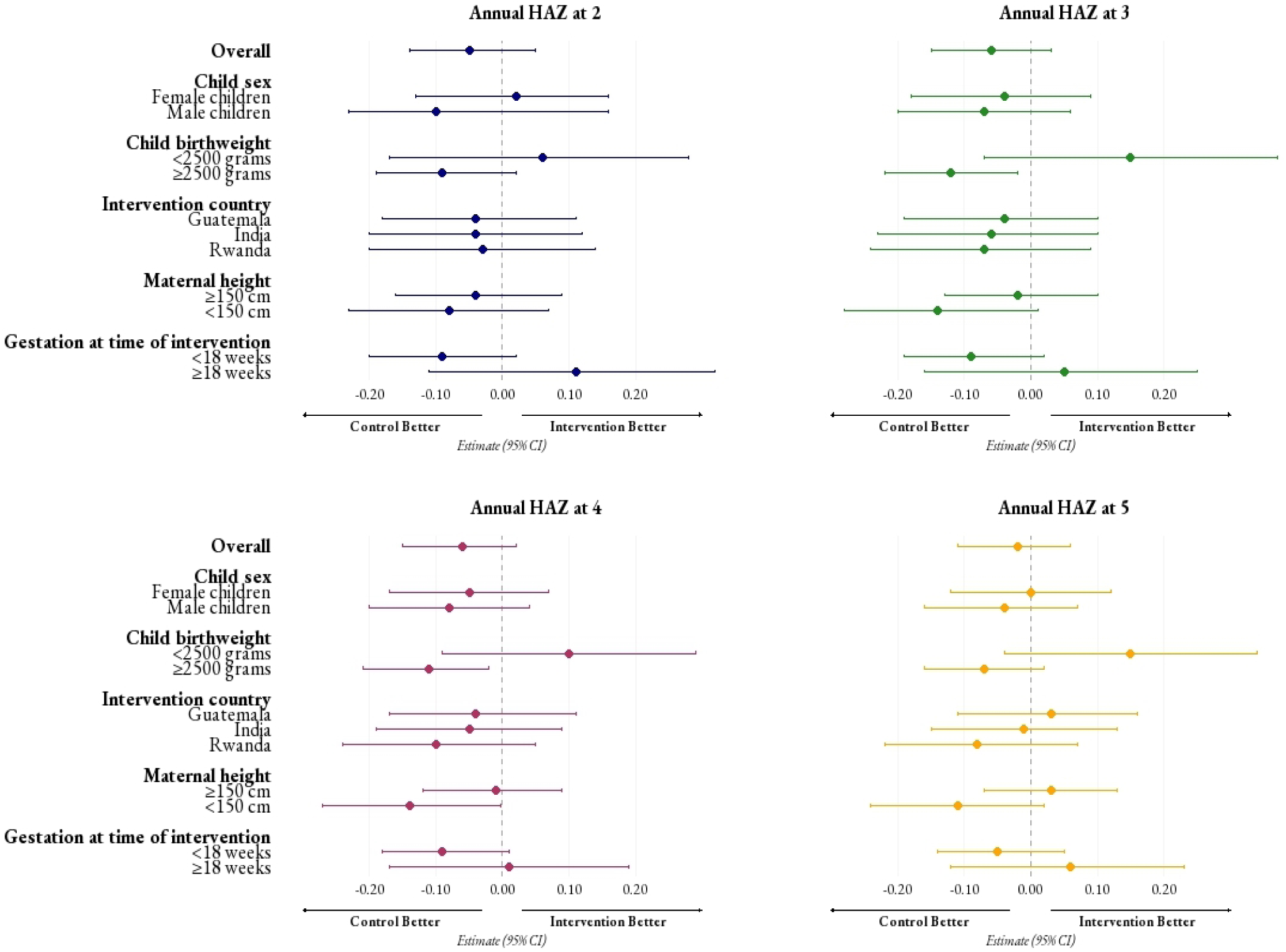
Subgroup analyses of the effects of the intervention on annual HAZ.

**Figure 4:**
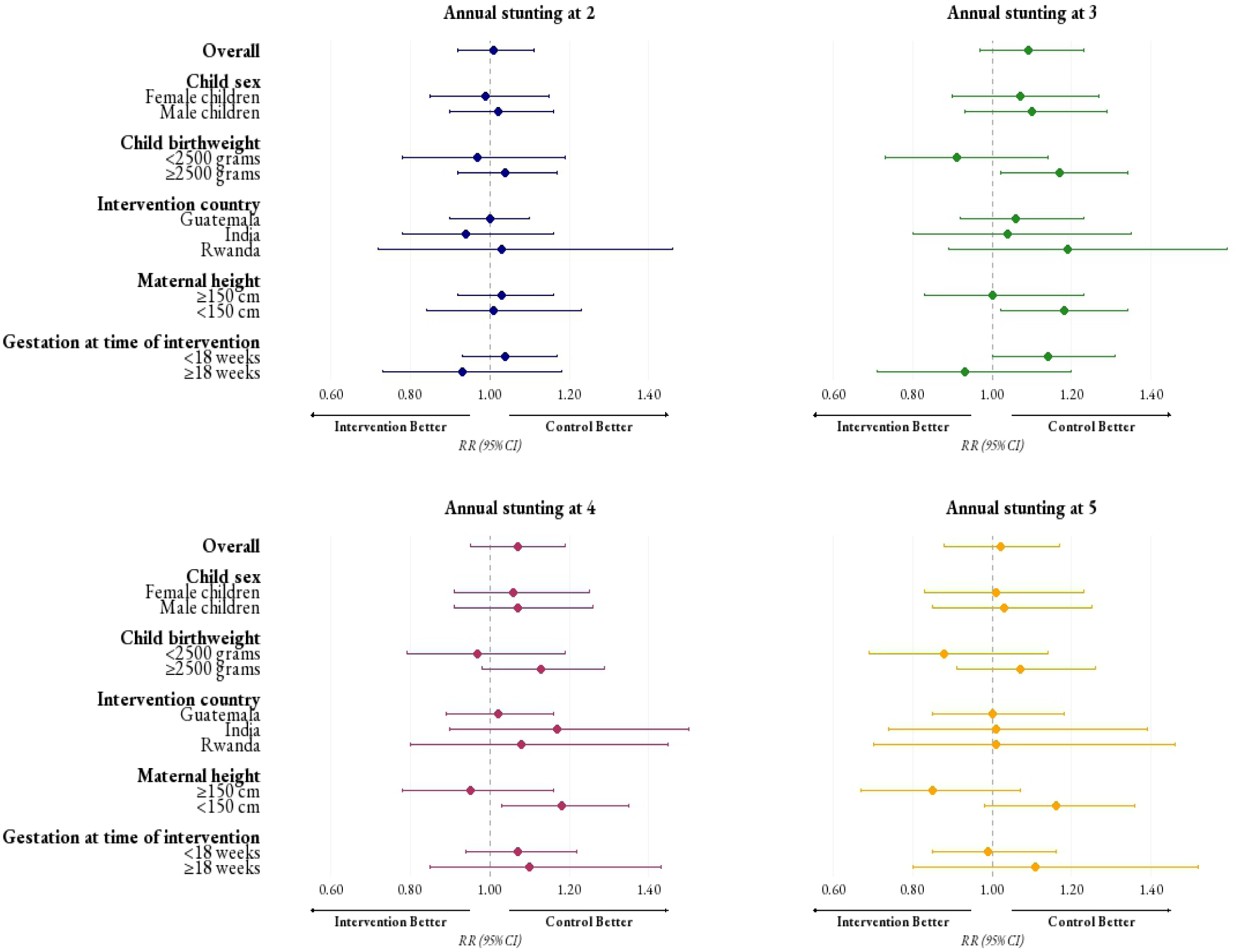
Subgroup analyses of the effects of the intervention on annual stunting.

Shown is the analysis of the effects of the intervention on HAZ at 2 to 5 years of age in the overall sample and in prespecified subgroups. Child sex was a binary variable and collected at birth, child birthweight was originally measured in grams at birth, which was categorized as <2500 grams [low birthweight as per WHO definition] and ≥ 2500 grams [normal birthweight as per WHO definition], intervention country is a categorical variable which represents the country of the intervention, maternal height was measured in centimeters at baseline [before randomization] which was categorized <150 cm [short – as per WHO cut-off] versus ≥150 cm [normal height as per WHO cut-off] and gestation at the time of intervention refers to the time at which the women in the intervention group received an LPG cookstove. A 95 % confidence interval was used for the total sample included in all linear regression models.

Similarly, the subgroup analyses on annual stunting models showed no subgroup appeared to be benefited by the intervention (detail results are provided in the supplementary file (S-table 8).

Shown is the analysis of the effects of the intervention on annual stunting at 2 to 5 years of age in the overall sample and in prespecified subgroups. Child sex was a binary variable and collected at birth, child birthweight was originally measured in grams at birth, which was categorized as <2500 grams [low birthweight as per WHO definition] and ≥ 2500 grams [normal birthweight as per WHO definition], intervention country is a categorical variable which represents the country of the intervention, maternal height was measured in centimeters at baseline [before randomization] which was categorized <150 cm [short – as per WHO cut-off] versus ≥150 cm [normal height as per WHO cut-off] and gestation at the time of intervention refers to the time at which the women in the intervention group received an LPG cookstove. A 95 % confidence interval was used for the total sample included in all log-binomial regression models.

## Discussion

This is the first known study to evaluate the effect of an LPG cookstove and clean fuel intervention, implemented during gestation and infancy, on child linear growth outcomes, including HAZ, stunting, and severe stunting assessed between 2 and 5 years of age. While the intervention was nearly exclusively used and thus able to markedly reduce the gestational and <12 months of age exposure to PM_2.5_, CO, and BC in the intervention households compared to the control households, the intervention did not improve HAZ between ages 2 and 5. We also found no change in the relative risk of stunting or severe stunting between the intervention and control groups or in subgroups.

These findings are in contrast with observational studies showing an association between HAP and linear growth in different LMICs (17, 18, 20). While such studies frequently propose possible biological explanations, such as inflammation, infection, or nutritional impacts, these mechanisms remain hypothetical and are largely inferred rather than clinically demonstrated. More importantly, these studies capture associations but cannot establish whether HAP directly affects linear growth, as they are observational in nature and therefore susceptible to residual confounding and bias (37). In contrast, randomized trials like HAPIN reduce confounding and allow a more direct assessment of whether reducing the exposure of HAP improves linear growth. Despite substantial reductions in HAP through LPG use, no effect was observed on HAZ or stunting, suggesting that HAP reduction alone is unlikely to alter growth, particularly in settings where multiple nutritional, infectious, and socioeconomic determinants of growth coexist [Kalam et al., forthcoming]. These results align with earlier findings that the HAPIN intervention had no effect on fetal growth, birthweight, and stunting at one year (22, 25, 26). These are well-known predictors of growth faltering and stunting.

There are several possible reasons for this finding. Firstly, as has been noted, most evidence suggests that impaired linear growth is largely established by age 2, and that early interventions are unlikely to have an effect after age 2 (4, 38, 39). In this respect, our findings, though novel in that they investigate effects of an HAP intervention that was effective in reducing the target exposure, are consistent with the literature on HAZ and stunting through age 5 (8). Importantly, the intervention ended when children reached at 12 months, and thereafter most households resumed using biomass fuels for cooking, exposing them to the substantial levels of HAP similar to those were experienced pre-trial and by the control group. Our aim was to measure whether reducing household air pollution during the gestational period and the child’s first year of life, during which growth is thought to be most sensitive, would have a sustained impact on linear growth through early childhood, up to age 5. By following children beyond the intervention period, we sought to assess whether early-life exposure reduction could translate into measurable improvements in HAZ, even after households largely returned to biomass fuel use.

Secondly, in many low-resource settings, children may continue to be exposed to PM_2.5_ from community-level emissions from neighboring households still using biomass fuels, from ambient air pollution, or from other local sources such as trash burning and agricultural fires (40–43). These community-level and outdoor sources contribute to background levels of PM_2.5_ that are often well above WHO guidelines, potentially offsetting the benefits achieved within the home (44–46). Although the intervention reduced personal PM_2.5_ exposure, measured exposures remained above the value recommended by the WHO annual guideline (5 μg/m³) (47). Greater reductions in exposure, bringing concentrations closer to the WHO guideline, may therefore be needed to produce measurable improvements in HAZ. Similarly, CO may still be present at low levels from intermittent sources such as kerosene lamps, motor vehicles, or secondary use of biomass for tasks like water heating or burning trash. Even modest CO exposure can interfere with oxygen delivery and metabolism in young children (48, 49). Black carbon, on the other hand, is closely linked with both household and ambient biomass burning. While LPG use drastically reduces BC emissions and exposures from cooking, neighborhood-level emissions can remain high in areas where biomass continues to be used by others. Black carbon is associated with oxidative stress and systemic inflammation, which are potential pathways influencing impaired child growth (9). Moreover, the biological effects of air pollution on growth, including chronic systemic inflammation, altered gut microbiota, endocrine disruption, and reduced nutrient absorption, may require prolonged and substantial exposure reduction to produce measurable changes in anthropometry (10–12). If residual exposure to PM_2.5_, CO, and BC remained above harmful thresholds, the physiological benefits necessary for supporting linear growth may not have materialized. However, in the HAPIN setting, ambient air pollution was likely minor compared with household sources (50).

Beyond exposure to air pollution, child linear growth is a complex, multifactorial process driven by interrelated factors including chronic undernutrition, recurrent infections, systemic inflammation, poor caregiving practices, and environmental exposures (38, 51). Although our intervention reduced exposures to HAP during early life, it did not directly address dietary intake. Persistent macronutrient deficiencies, limited dietary diversity, and inadequate micronutrient intake (e.g., iron, zinc, vitamin A) likely continued to constrain growth potential in this population (7, 52, 53). Without concurrent nutritional interventions, children may have lacked the necessary substrates and cofactors for tissue growth, even in a cleaner air environment (54, 55). Moreover, the nutrition of mothers before and during pregnancy plays a pivotal role in determining fetal growth and size at birth (39). Inadequate maternal nutrition, which the intervention did not address, may have led to intrauterine growth restriction—a key predictor of stunting in early life. While HAP has been hypothesized to contribute to impaired growth, it is only one component of a broader risk environment. Its independent effect may be modest, particularly when not addressed in tandem with other key drivers of stunting.

Additionally, evidence suggests that single-sector interventions are often insufficient to fully address growth faltering. For example, sanitation-only interventions generally have minimal effects on linear growth (56). Maternal and child nutrition interventions, including micronutrient supplementation, fortified foods, and conditional cash transfers, produce modest gains while behavior change and education programs (e.g., complementary feeding education) and food-based interventions (fortified blended foods, lipid-based supplements, animal-source foods) achieve moderate improvements (55, 57, 58). Integrated programs combining nutrition, education, and deworming yield the largest gains (59). These findings help contextualize our results, suggesting that reducing household air pollution alone, without concurrent improvements in other risk factors, including nutrition, infection control, and caregiving practices, is unlikely to produce meaningful improvements in linear growth.

Lastly, physiological benefits from improved environmental exposures may take time to manifest in measurable anthropometric changes (60, 61). It is possible that reductions in air pollution improved intermediate outcomes such as systemic inflammation or immune function (62), but these changes may not have translated into linear growth gains within five years. Catch-up growth, particularly for children who were already stunted, may require multi-pronged interventions that go beyond environmental health, including nutritional supplementation, infection control, and developmental stimulation (27).

Despite offering rigorous study design, rich longitudinal follow-up on child HAZ, air pollutant exposure data, and an adequate sample from a multi-country setting, there were several limitations that affect how the presented results should be interpreted. Firstly, the missingness during the extended follow-up: a significant proportion of infants had missing or invalid infancy exposure measures, missing anthropometric data at follow-up visits, and implausible height or HAZ measurements at different follow-up visits. Although the extended follow-up was less intensive, as compared to the primary follow-up during the intervention, the missingness could influence the estimates and potentially bias the results. Additionally, exposure measurement during the gestational period was largely impacted by the COVID-19 induced lockdown and mobility restrictions. During infancy, we had substantial missingness on child’s exposure data due to operational and logistical issues. Secondly, reliance on self-reported covariates may have introduced misclassification due to recall error or social desirability bias. Such misclassification could result in residual confounding if important covariates were not measured accurately, potentially attenuating or obscuring true associations between exposures, interventions, and child growth outcomes. Thirdly, while the multi-country design enhances contextual relevance, the study populations may not be representative of broader populations within participating countries. Lastly, with respect to intervention timing, the HAPIN trial did not capture the earliest stages of conception and fetal development. Although the intervention demonstrated high fidelity and adherence, it was implemented primarily after the first trimester of gestation and therefore may have missed critical windows of early growth (63, 64).

## Conclusion

We observed significant reductions in PM_2.5_, CO and BC exposures during gestation and PM_2.5_, and CO during infancy in the intervention group. However, these benefits did not translate into improving HAZ or reducing the risk of stunting at ages 2 through 5 years. Although HAZ increased over time, with a corresponding decline in stunting prevalence, these gains were not attributable to the LPG cookstove and clean fuel intervention in this multi-country cohort. Hence, we do not support using LPG cookstoves with clean fuel during gestation and infancy as a strategy to improving HAZ or reducing the risk of stunting through 2 to 5 years of age. Exposure–response analyses may offer valuable complementary evidence to contextualize these findings.

## Data Availability

The trial protocol (including planned primary and secondary outcomes), informed consent form, and primary outcome statistical analysis plans can be accessed at https://clinicaltrials.gov/study/NCT02944682. De-identified data associated with these analyses will be available through the Emory University Dataverse repository, linked to the DOI of this publication (https://doi.org/10.15139/S3/4WGVHC, Dataverse Link: https://dataverse.unc.edu/dataset.xhtml?persistentId=doi:10.15139/S3/4WGVHC. Other materials can be accessed upon request.

## Acknowledgement

The investigators would like to thank the members of the advisory committee – Drs. Patrick Breysse, Donna Spiegelman, and Joel Kaufman -for their valuable insight and guidance throughout the implementation of the trial. We also wish to acknowledge all research staff and study participants for their dedication to and participation in this important trial.

A multidisciplinary, independent Data and Safety Monitoring Board (DSMB) appointed by the National Heart, Lung, and Blood Institute (NHLBI) monitored the quality of the data and protected the safety of patients enrolled in the HAPIN trial. The DSMB consisted of: Catherine Karr (Chair), Nancy R. Cook, Stephen Hecht, Joseph Millum, Nalini Sathiakumar (deceased), Paul K. Whelton, and Gail Weinmann and Thomas Croxton (Executive Secretaries). Program Coordination: Gail Rodgers, Bill & Melinda Gates Foundation; Claudia L. Thompson, National Institute of Environmental Health Sciences; Mark J. Parascandola, National Cancer Institute; Marion Koso-Thomas, Eunice Kennedy Shriver National Institute of Child Health and Human Development; Joshua P. Rosenthal, Fogarty International Center; Concepcion R. Nierras, NIH Office of Strategic Coordination – The Common Fund; Katherine Kavounis, Dong-Yun Kim, Barry S. Schmetter (deceased), and Antonello Punturieri, NHLBI.

This research represents the NIH’s contribution to the Global Alliance for Chronic Diseases (GACD) coordinated call for research on prevention and management of chronic lung diseases for 2016.

The findings and conclusions in this report are those of the authors alone and do not necessarily represent the official position of the US National Institutes of Health, US Department of Health and Human Services, or the Gates Foundation. It is subject to the NIH Public Access Policy. Through acceptance of this federal funding, NIH has been given a right to make this manuscript publicly available in PubMed Central upon the Official Date of Publication, as defined by NIH. Under the grant conditions of the Gates Foundation, a Creative Commons Attribution 4.0 License has already been assigned to the Author Accepted Manuscript version that might arise from this submission.

## Supporting information

S-box 1: Statistical analyses plan

S-table 1: Number of children participated in all follow-up visits

S-table 2: Number of children participated in any follow-up visits

S-table 3: Baseline household and maternal characteristics of the missing sample

S-table 5: Child dietary diversity score

S-table 6. Full Mixed-Effects Model Results

S-table 7: Subgroup analyses on the effect of the intervention in different sub-groups on HAZ at ages 2, 3, 4 and 5

S-table 8: Subgroup analyses on the effect of the intervention in different sub-groups on stunting at ages 2, 3, 4 and 5

## References

1. Organization WH. Levels and trends in child malnutrition child malnutrition: UNICEF/WHO/World Bank Group Joint Child Malnutrition Estimates: Key findings of the 2023 edition: World Health Organization; 2023.

2. De Sanctis V, Soliman A, Alaaraj N, Ahmed S, Alyafei F, Hamed N. Early and Long-term Consequences of Nutritional Stunting: From Childhood to Adulthood. Acta Biomed. 2021;92(1):e2021168.

3. McGovern ME, Krishna A, Aguayo VM, Subramanian SV. A review of the evidence linking child stunting to economic outcomes. Int J Epidemiol. 2017;46(4):1171–91.

4. Georgiadis A, Penny ME. Child undernutrition: opportunities beyond the first 1000 days. The Lancet Public Health. 2017;2(9):e399.

5. Bhargava A. Protein and micronutrient intakes are associated with child growth and morbidity from infancy to adulthood in the Philippines. The Journal of nutrition. 2016;146(1):133–41.

6. Benjamin-Chung J, Mertens A, Colford Jr JM, Hubbard AE, van der Laan MJ, Coyle J, et al. Early-childhood linear growth faltering in low-and middle-income countries. Nature. 2023;621(7979):550–7.

7. Li Z, Kim R, Vollmer S, Subramanian SV. Factors Associated With Child Stunting, Wasting, and Underweight in 35 Low-and Middle-Income Countries. JAMA Netw Open. 2020;3(4):e203386.

8. Johnson NM, Hoffmann AR, Behlen JC, Lau C, Pendleton D, Harvey N, et al. Air pollution and children’s health-a review of adverse effects associated with prenatal exposure from fine to ultrafine particulate matter. Environ Health Prev Med. 2021;26(1):72.

9. Sinharoy SS, Clasen T, Martorell R. Air pollution and stunting: a missing link? Lancet Glob Health. 2020;8(4):e472–e5.

10. Ladd-Acosta C, Feinberg JI, Brown SC, Lurmann FW, Croen LA, Hertz-Picciotto I, et al. Epigenetic marks of prenatal air pollution exposure found in multiple tissues relevant for child health. Environ Int. 2019;126:363–76.

11. Iodice S, Hoxha M, Ferrari L, Carbone IF, Anceschi C, Miragoli M, et al. Particulate Air Pollution, Blood Mitochondrial DNA Copy Number, and Telomere Length in Mothers in the First Trimester of Pregnancy: Effects on Fetal Growth. Oxid Med Cell Longev. 2018;2018:5162905.

12. Burris HH, Baccarelli AA. Air pollution and in utero programming of poor fetal growth. Epigenomics. 2017;9(3):213–6.

13. Sania A, Sudfeld CR, Danaei G, Fink G, McCoy DC, Zhu Z, et al. Early life risk factors of motor, cognitive and language development: a pooled analysis of studies from low/middle-income countries. BMJ Open. 2019;9(10):e026449.

14. Gladstone M, Oliver C, Van den Broek N. Survival, morbidity, growth and developmental delay for babies born preterm in low and middle income countries -a systematic review of outcomes measured. PLoS One. 2015;10(3):e0120566.

15. Christian P, Murray-Kolb LE, Tielsch JM, Katz J, LeClerq SC, Khatry SK. Associations between preterm birth, small-for-gestational age, and neonatal morbidity and cognitive function among school-age children in Nepal. BMC Pediatr. 2014;14:58.

16. Roomaney RA, Wright CY, Cairncross E, Abdelatif N, Cois A, Turawa EB, et al. Estimating the burden of disease attributable to household air pollution from cooking with solid fuels in South Africa for 2000, 2006 and 2012. S Afr Med J. 2022;112(8b):718–28.

17. Islam S, Rana MJ, Mohanty SK. Cooking, smoking, and stunting: Effects of household air pollution sources on childhood growth in India. Indoor Air. 2021;31(1):229–49.

18. deSouza PN, Hammer M, Anthamatten P, Kinney PL, Kim R, Subramanian SV, et al. Impact of air pollution on stunting among children in Africa. Environ Health. 2022;21(1):128.

19. Caulfield LE, Richard SA, Rivera JA, Musgrove P, Black RE. Stunting, wasting, and micronutrient deficiency disorders. Disease Control Priorities in Developing Countries 2nd edition. 2006.

20. Caleyachetty R, Lufumpa N, Kumar N, Mohammed NI, Bekele H, Kurmi O, et al. Exposure to household air pollution from solid cookfuels and childhood stunting: a population-based, cross-sectional study of half a million children in low- and middle-income countries. Int Health. 2022;14(6):639–47.

21. Bickton FM, Ndeketa L, Sibande GT, Nkeramahame J, Payesa C, Milanzi EB. Household air pollution and under-five mortality in sub-Saharan Africa: an analysis of 14 demographic and health surveys. Environ Health Prev Med. 2020;25(1):67.

22. Checkley W, Thompson LM, Sinharoy SS, Hossen S, Moulton LH, Chang HH, et al. Effects of Cooking with Liquefied Petroleum Gas or Biomass on Stunting in Infants. N Engl J Med. 2024;390(1):44–54.

23. Boamah-Kaali E, Jack DW, Ae-Ngibise KA, Quinn A, Kaali S, Dubowski K, et al. Prenatal and Postnatal Household Air Pollution Exposure and Infant Growth Trajectories: Evidence from a Rural Ghanaian Pregnancy Cohort. Environ Health Perspect. 2021;129(11):117009.

24. Dutta A, Alexander D, Karrison T, Morhasson-Bello O, Wilson N, Atalabi OM, et al. Household air pollution, ultrasound measurement, fetal biometric parameters and intrauterine growth restriction. Environ Health. 2021;20(1):74.

25. Clasen TF, Chang HH, Thompson LM, Kirby MA, Balakrishnan K, Diaz-Artiga A, et al. Liquefied Petroleum Gas or Biomass for Cooking and Effects on Birth Weight. N Engl J Med. 2022;387(19):1735–46.

26. Checkley W, Thompson LM, Hossen S, Nicolaou L, Williams KN, Hartinger SM, et al. Cooking with liquefied petroleum gas or biomass and fetal growth outcomes: a multi-country randomised controlled trial. Lancet Glob Health. 2024;12(5):e815–e25.

27. Draper CE, Yousafzai AK, McCoy DC, Cuartas J, Obradović J, Bhopal S, et al. The next 1000 days: building on early investments for the health and development of young children. The Lancet. 2024;404(10467):2094–116.

28. Lancet T. The next 1000 days: the forgotten ages of child health. 2024. p. 2021.

29. Clasen T, Checkley W, Peel JL, Balakrishnan K, McCracken JP, Rosa G, et al. Design and Rationale of the HAPIN Study: A Multicountry Randomized Controlled Trial to Assess the Effect of Liquefied Petroleum Gas Stove and Continuous Fuel Distribution. Environ Health Perspect. 2020;128(4):47008.

30. Quinn AK, Williams KN, Thompson LM, Harvey SA, Piedrahita R, Wang J, et al. Fidelity and adherence to a liquefied petroleum gas stove and fuel intervention during gestation: the multi-country Household Air Pollution Intervention Network (HAPIN) randomized controlled trial. International journal of environmental research and public health. 2021;18(23):12592.

31. Quinn AK, Williams K, Thompson LM, Rosa G, Diaz-Artiga A, Thangavel G, et al. Compensating control participants when the intervention is of significant value: experience in Guatemala, India, Peru and Rwanda. BMJ Glob Health. 2019;4(4):e001567.

32. Butte NF, Garza C, de Onis M. Evaluation of the feasibility of international growth standards for school-aged children and adolescents. J Nutr. 2007;137(1):153–7.

33. De Onis M. WHO Child Growth Standards Length/height-for-age, weight-for-age, weight-for-length, weight-for-height and body mass index-for-age. Methods and development. 2006.

34. Johnson MA, Steenland K, Piedrahita R, Clark ML, Pillarisetti A, Balakrishnan K, et al. Air Pollutant Exposure and Stove Use Assessment Methods for the Household Air Pollution Intervention Network (HAPIN) Trial. Environ Health Perspect. 2020;128(4):47009.

35. Group WHOMGRS. WHO Child Growth Standards based on length/height, weight and age. Acta Paediatr Suppl. 2006;450:76–85.

36. Fund UNCs, Organization WH. UNICEF/WHO Low Birthweight Estimates: levels and trends 2000-2020: World Health Organization; 2023.

37. Bruce NG, Dherani MK, Das JK, Balakrishnan K, Adair-Rohani H, Bhutta ZA, et al. Control of household air pollution for child survival: estimates for intervention impacts. BMC Public Health. 2013;13 Suppl 3(Suppl 3):S8.

38. Bhutta ZA, Das JK, Rizvi A, Gaffey MF, Walker N, Horton S, et al. Evidence-based interventions for improvement of maternal and child nutrition: what can be done and at what cost? Lancet. 2013;382(9890):452–77.

39. Mertens A, Benjamin-Chung J, Colford Jr JM, Coyle J, Van der Laan MJ, Hubbard AE, et al. Causes and consequences of child growth faltering in low-resource settings. Nature. 2023;621(7979):568–76.

40. Alter NC, Whitman EM, Bellinger DC, Landrigan PJ. Quantifying the association between PM(2.5) air pollution and IQ loss in children: a systematic review and meta-analysis. Environ Health. 2024;23(1):101.

41. Diseases GBD, Injuries C. Global burden of 369 diseases and injuries in 204 countries and territories, 1990-2019: a systematic analysis for the Global Burden of Disease Study 2019. Lancet. 2020;396(10258):1204–22.

42. Wang JG, Bose S, Viegi G. Reduced Childhood Ambient Fine Particulate Matter Exposure and Subsequent Asthma Incidence: How Low Is Low Enough? Ann Am Thorac Soc. 2024;21(10):1389–90.

43. Zhu K, Kawyn MN, Kordas K, Mu L, Yoo EH, Seibert R, et al. Assessing exposure to household air pollution in children under five: A scoping review. Environ Pollut. 2022;311:119917.

44. Farmer SA, Nelin TD, Falvo MJ, Wold LE. Ambient and household air pollution: complex triggers of disease. Am J Physiol Heart Circ Physiol. 2014;307(4):H467–76.

45. Feng J, Wang Y, Meng X, Fang Y. Global trends in disease burdens attributable to ambient and household air pollution: a comparative study of ten populous countries. Front Public Health. 2025;13:1629616.

46. Song C, Lim CC, Gurmu BL, Kim M, Lee S, Park J, et al. Comparison of Personal or Indoor PM(2.5) Exposure Level to That of Outdoor: Over Four Seasons in Selected Urban, Industrial, and Rural Areas of South Korea: (K-IOP Study). Int J Environ Res Public Health. 2023;20(17).

47. Organization WH. WHO ambient air quality database, 2022 update: status report: World Health Organization; 2023.

48. Lewis A, Forti RM, Alomaja O, Mesaros C, Piel S, Greenwood JC, et al. Preliminary Research: Application of Non-Invasive Measure of Cytochrome c Oxidase Redox States and Mitochondrial Function in a Porcine Model of Carbon Monoxide Poisoning. J Med Toxicol. 2022;18(3):214–22.

49. Wen T, Liang J, Wei Y, Lin W, Pan L. Analysis of prognosis of neurological sequelae in children with carbon monoxide poisoning. Sci Rep. 2024;14(1):29972.

50. Liao J, Kirby MA, Pillarisetti A, Piedrahita R, Balakrishnan K, Sambandam S, et al. LPG stove and fuel intervention among pregnant women reduce fine particle air pollution exposures in three countries: Pilot results from the HAPIN trial. Environ Pollut. 2021;291:118198.

51. Escher NA, Andrade GC, Ghosh-Jerath S, Millett C, Seferidi P. The effect of nutrition-specific and nutrition-sensitive interventions on the double burden of malnutrition in low-income and middle-income countries: a systematic review. The Lancet Global Health. 2024;12(3):e419–e32.

52. Ahmed KY, Dadi AF, Ogbo FA, Page A, Agho KE, Akalu TY, et al. Population-modifiable risk factors associated with childhood stunting in sub-Saharan Africa. JAMA network open. 2023;6(10):e2338321-e.

53. Nguyen PH, Scott S, Neupane S, Tran LM, Menon P. Social, biological, and programmatic factors linking adolescent pregnancy and early childhood undernutrition: a path analysis of India’s 2016 National Family and Health Survey. Lancet Child Adolesc Health. 2019;3(7):463–73.

54. Coll-Seck A, Clark H, Bahl R, Peterson S, Costello A, Lucas T. Framing an agenda for children thriving in the SDG era: a WHO-UNICEF-Lancet Commission on Child Health and Wellbeing. Lancet. 2019;393(10167):109–12.

55. Keats EC, Das JK, Salam RA, Lassi ZS, Imdad A, Black RE, et al. Effective interventions to address maternal and child malnutrition: an update of the evidence. Lancet Child Adolesc Health. 2021;5(5):367–84.

56. Freeman MC, Garn JV, Sclar GD, Boisson S, Medlicott K, Alexander KT, et al. The impact of sanitation on infectious disease and nutritional status: A systematic review and meta-analysis. International journal of hygiene and environmental health. 2017;220(6):928–49.

57. Hossain M, Choudhury N, Abdullah KAB, Mondal P, Jackson AA, Walson J, et al. Evidence-based approaches to childhood stunting in low and middle income countries: a systematic review. Archives of Disease in Childhood. 2017;102(10):903–9.

58. Sharma IK, Di Prima S, Essink D, Broerse JE. Nutrition-sensitive agriculture: a systematic review of impact pathways to nutrition outcomes. Advances in Nutrition. 2021;12(1):251–75.

59. Ghodsi D, Omidvar N, Nikooyeh B, Roustaee R, Shakibazadeh E, Al-Jawaldeh A. Effectiveness of community nutrition-specific interventions on improving malnutrition of children under 5 years of age in the eastern mediterranean region: A systematic review and meta-analysis. International Journal of Environmental Research and Public Health. 2021;18(15):7844.

60. He S, Lundberg B, Hallberg J, Klevebro S, Pershagen G, Eneroth K, et al. Joint association of air pollution exposure and inflammation-related proteins in relation to infant lung function. Int J Hyg Environ Health. 2024;255:114294.

61. Tseng W-J, Lu J-H, Chao H-R, Tsai M-H, Chang Y-T, Wang L-J, et al. Associations between children’s exposure to PM2. 5 and their serum inflammatory responses in Taiwan. Aerosol and air quality research. 2022;22(12):220288.

62. Wittkopp S, Walzer D, Thorpe L, Roberts T, Xia Y, Gordon T, et al. Portable air cleaner use and biomarkers of inflammation: A systematic review and meta-analysis. American heart journal plus: cardiology research and practice. 2022;18:100182.

63. Rosofsky AS, Fabian MP, Ettinger de Cuba S, Sandel M, Coleman S, Levy JI, et al. Prenatal ambient particulate matter exposure and longitudinal weight growth trajectories in early childhood. International journal of environmental research and public health. 2020;17(4):1444.

64. von Salmuth V, Brennan E, Kerac M, McGrath M, Frison S, Lelijveld N. Maternal-focused interventions to improve infant growth and nutritional status in low-middle income countries: A systematic review of reviews. PloS one. 2021;16(8):e0256188.

